# Lung injury in patients with or suspected COVID-19 : a comparison between lung ultrasound and chest CT-scanner severity assessments, an observational study

**DOI:** 10.1101/2020.04.24.20069633

**Authors:** Mehdi Benchoufi, Jerôme Bokobza, Anthony Chauvin, Elisabeth Dion, Marie-Laure Baranne, Fabien Levan, Maxime Gautier, Delphine Cantin, Thomas d’Humières, Cédric Gil-Jardiné, Sylvain Benenati, Mathieu Orbelin, Mikaël Martinez, Nathalie Pierre-Kahn, Abdourahmane Diallo, Eric Vicaut, Pierre Bourrier

**Affiliations:** Center for Clinical Epidemiology, Hôtel-Dieu Hospital, Assistance Publique-Hôpitaux de Paris (AP-HP), Paris, France; METHODS Team, Center for Research in Epidemiology and Statistics Sorbonne Paris Cité (CRESS- UMR 1153), Paris, France; Adult Emergency Department, Hôpital Cochin, Assistance Publique Hôpitaux de Paris, Paris, France; Adult Emergency Department, Hôpital Lariboisière, Assistance Publique Hôpitaux de Paris, University Diderot, Paris, France; Imaging department Hôtel Dieu, Assistance Publique-Hôpitaux de Paris, Université de Paris, Paris, France; INSERM U1149, Centre de Recherche de l’Inflammation (CRI), Paris, France; Adult Emergency Department, Hôpital Lariboisière, Assistance Publique Hôpitaux de Paris, Paris, France; Physiology department, Henri Mondor University Hospital, APHP, Créteil, France; Adult Emergency Department SAMU-SMUR, Pellegrin Hospital, University Hospital Center, Bordeaux, France; Bordeaux Population Health, INSERM U1219, IETO Team, Bordeaux University, Bordeaux, France; Adult Emergency Department, Hospital Group South Ile-de-France, Melun, France; Adult Emergency Department, New Civil Hospital, Strasbourg, France; Adult Emergency Department, Forez Hospital Center, Montbrison, France; Nord Emergency Network Ligérien Ardèche (REULIAN), Hospital Center Le Corbusier, Firminy, France; Clinical Trial Unit Hospital, Lariboisière St-Louis AP-HP, Paris University; Imaging Department Saint-Louis Hospital, Assistance Publique des Hôpitaux de Paris (AP-HP), Paris, France

**Keywords:** COVID-19, Pulmonary ultrasound, Screening, Diagnosis improvement, Chest-CT

## Abstract

**Background:** Chest CT (CT) is the reference for assessing pulmonary injury in suspected or diagnosed COVID-19 with signs of clinical severity. We explored the role of lung ultrasonography (LU) in quickly assessing lung status in these patients.

**Methods:** eChoVid is a multicentric study based on routinely collected data, conducted in 3 emergency units of Assistance Publique des Hôpitaux de Paris (APHP); 107 patients were included between March 19, 2020 and April 01, 2020 and underwent LU, a short clinical assessment by 2 emergency physicians blinded to each other’s and a CT. LU consisted of scoring lesions in 8 chest zones from 0 to 3, defining a severity global score (GS) ranging from 0 to 24. CT severity score ranged from 0 to 3 according to the extent of interstitial pneumonia signs. 48 patients underwent LU by both an expert and a newly trained physician.

**Findings:** The GS showed good performance to predict CT severity assessment of COVID-19 as normal versus pathologic: AUC=0.93, maximal Youden index 1 with 95% sensitivity, and 83% specificity. Similar performance was found for CT assessment as normal or minimal versus moderate or severe (n=90): AUC 0.89, maximal Youden index 7 with 86% sensitivity, and 78% specificity. Good agreement was found for zone scoring assessed by new trainee (30mn theory + 30mn practice) and expert (n=14,14*8 checkpoints), weighted kappa 0.85-1; moderate agreement was found for new trainee (n=48, 30mn theory) and expert, kappa 0.62-0.81.

**Interpretation:** GS score is a simple tool to assess lung damage severity in patients with suspected or diagnosed COVID-19. Comparing the performance of new trainees and expert physicians opens a path for adoption beyond the scope of experts. LU is a good candidate for patients triage, especially in case of CT availability issues.

## Introduction

Since December 2019, the SARS-CoV2 epidemic spread from Wuhan, Hubei Province, China [1]. In May 11, 2020, WHO declared that SARS-Cov2 infection (COVID-19) was a pandemic [2]. First symptoms are fever, fatigue, dry cough, and dyspnea [3]. Pneumonia associated with COVID-19 is foremost in the symptomatology and prognosis [4]. Respiratory symptoms ranges from discrete to severe dyspnea signalling the evolution to acute respiratory distress syndrome (ARDS) [4].

Hence, we need to quickly discriminate patients that could eventually be sent back home and those who deserve special attention and whose remote follow-up or hospitalised care must be organized. In the context of COVID-19 management French recommendations, CT is the reference to assess pulmonary involvment status in suspected or diagnosed COVID-19 with signs of clinical severity [5–7] and to infer screening and orientation decisions.

The major CT signs in Covid-19 pneumonia are the following: frequent bilateral lung involvement, multiple mottling, ground-glass opacities, crazy-paving lesions [8]. The US and French radiology societies have published guidelines to limit or rule out chest X-rays for managing COVID-19 [7], CT being the gold standard assessment [5] and even for some authors to diagnose advanced SARS-CoV2 infection because of its highest sensitivity [9].

However, CT being radiating, limited by availability issues given the heavy patient flow in hospitals in the COVID-19 outbreak or the absence of CT in some hospital and medical centers, and by transportation issues for hemodynamically unstable patients, alternative solutions to assess and screen affected or suspected COVID-19 patients are needed. Lung ultrasonography (LU) may be a good candidate. Some evidence suggests that interstitial pneumonia signs (B-lines), sub-pleural condensations (wedge signs) and foci of consolidations (hepatization) can be detected with LU [10]. Moreover, LU has many advantages: carried out in a few minutes, at the patient’s bedside; interpretable in real-time results; interpreted by the doctor in charge, non-radiating, and relatively low-cost (especially handheld devices).

Today, the use of LU is framed by relatively generic guidelines [11] and, to our knowledge, no study has precisely established its place in affected or suspected COVID-19 management. Recent feedback from Chinese and Italian teams using LU as a quick severity assessment tool for pneumonia and ARDS; as a follow-up tool; or even as an early diagnosis tool [10, 12] suggest that LU could be useful in distinguishing patients with affected or suspected COVID-19, those without particular grounds for concern and those to be referred to intensive management. LU could be compared to CT for assessing lung damages severity and thus infer insights on how LU could be used as a triage tool.

The primary objective of this observational multicentric study based on routinely collected data was to assess the concordance of the evaluation of lung damage severity by LU and chest CT reports in patients with suspected or diagnosed COVID-19 who had the 2 examinations at the same time. The secondary objective was to compare the performance of a newly trained operator and an expert operator in terms of ultrasound assessment of pulmonary lesions in suspected or diagnosed COVID-19 patients.

## Methods

### Study design and patient selection

This was a multicentric, observational non-randomized study, conducted in the emergency units (EU) of 3 hospitals of APHP: the EU of Lariboisière University Hospital and Cochin University Hospital and an EU located in Hôtel-Dieu Hospital, converted into a COVID-19 screening unit for APHP medical staff with suspected COVID-19.

Patients were included from March 19, 2020 to April 1, 2020. Inclusion criteria were age > 18 years with suspected or diagnosed COVID-19 who underwent CT. Exclusion criteria were patients for whom the LU exploration could not be performed (morbid obesity, extensive thoracic subcutaneous emphysema, absorbent subcutaneous infiltrations) or with any comorbidity that justified priority immediate intensive care.

### Data collection and data sources

After inclusion, each patient underwent both a clinical examination and LU, each by an emergency physician. Emergency physician and ultrasound operator were blinded to each other’s findings.

Some LU exams were conducted by 2 physicians: one expert, emergency or imaging physician and one physician newly trained in LU. The latter underwent a 30-min training protocol before proving able to explore normal lungs and to recognize lung abnormalities on ultrasound images from an image bank.

Clinical Data were collected during the physical examination and data related to the context were also collected (former patient journey, medical background, recent use of nonsteroidal anti-inflammatory drugs (NSAIDs) or long-term use in the context of a known pathology). We collected the result and date of RT-PCR COVID-19 test.

LU results of 8 fields (right antero-superior, left antero-superior, right antero-inferior, left antero-inferior, right postero-superior, left postero-superior, right postero-inferior and left postero-superior) and the operator level of expertise were collected following Table 1. CT results including signs of severity, in accordance with the recommendations of the French society of radiology, were extracted from the radiologist’s report::

◦ whether the lung injuries were typical or not of SARS-CoV2 infection.
◦ severity of lung injury ranging from minimal (up to 10% of involved pulmonary parenchyma), moderate (10%-25%), extended (25%-50%), severe (50%-75%), critical (>75%), as standardised by the French society of radiology [5].

Clinical data measurement tools were standard in the EU. We report the use of Ultrasonography available equipment with no specific requirement on machine performance: TE7 (Mindray), curved probe (2-5 MHz), Ulight (Sonoscanner), curved probe (2-5 MHz), E2EXP (Sonoscape), curved probe (3Mhz), Spark (Philips), curved probe (2-6 MHz), Vscan (General Electrics), linear probe (4-12 MHz).

### Variables

The primary outcome was the estimation of the agreement between lung damage severity as assessed by LU and chest CT reports.

For LU, we defined 4 grades of severity: 0, up to a maximum of 3 observed B-lines; 1, 4 to 8 B-lines, through intercostal space at one of the pulmonary bases; 2, B-lines in “curtain sign” (> 8 B-lines) and/or diffusion of more than 4 B-lines in two-thirds of the pulmonary field; 3, consolidation foci. Gradation was carried out for each pulmonary half-field in anterior and posterior, superior and inferior views (cf. Table 1).

The ultrasound score for assessing lung condition was derived from the standard LU score (LUS) [13, 14]. LUS is computed by checking 12 points on the upper and lower parts of anterior, lateral, and posterior regions of the left and right chest wall. We simplified this to 8 points on the upper and lower parts of the anterior and posterior regions of the left and right chest wall. Therefore, our total lung score ranged from 0 to 24 points.

For CT, 2 data points were collected from the radiology report: 1) the consistency of the lung lesions with SARS-Cov2 infection namely ground-glass areas or nodules, nodular or strip condensations, crazy paving and 2) 4 grades of severity according to volume of injured lung parenchyma volume: minimal (<10%), moderate (10-25%), severe (> 25%). We collapsed to “severe” the gradations “extended”, “severe”, “critical” of the French society of radiology.

The secondary outcome was to compare the performance of a new trainee physician and an ultrasonography expert. New trainees were taught with either a 30mn protocol of ultrasound theory with review of pathological images from an image bank and practice on a pool of 5 Covid suspected patients, either 30mn protocol of ultrasound theory with imaging review.

### Study size

A sample of size 90 patients with documented level of severity allows for estimating an AUC-ROC ≥ 85% with precision of ±5% or better.

### Statistical analysis

All quantitative data are summarized with mean ± SD or median (interquartile range). Qualitative data are summarized with number (%). We evaluated different methods to quantify LU and their ability to predict lung disease evaluation by chest CT:

1. the average score of the 8 quadrants was classified on an ordinal scale as (0 [0–1], [1–2], [2–3]); Weighted kappa was used to study agreement between ultrasound ordinal scale and quantification of severity by CT (as normal, minimal, moderate, severe);
2. a global score (GS) for LU was computed by adding individual score from the 8 quadrants. The performance of GS was evaluated by univariate logistic regression model using GS to predict CT disease quantification dichotomized as *normal* versus *pathologic, normal or minimal* versus *moderate or severe*, or *normal, minimal or moderate* versus *severe*. For each regression model, we calculated AUC, Brier score, and Youden’s index for the different values of the scores. Model validation for calibration and discrimination ability involved bootstrap replications, and degrees of optimism were calculated for C statistics and Brier score [15]. When comparing LU and CT severity scores (*normal or minimal* vs *moderate or severe*, or *normal, minimal or moderate* vs *severe*), patients data with collected CT status (pathological/normal) but with missing CT severity score were discarded.
3. A simple weighted ultrasound score (SWS) was calculated. We built a multivariate logistic regression model using the scores for each quadrant to predict the CT disease quantification. Then we built a simplified score by considering only the quadrants significantly associated with severity and rounding the coefficients of the logistic model to obtain an easily computable score. We checked the performance of SWS in a univariate logistic model to predict CT results as described above for the GS.

Some patients were evaluated for LU by both an expert and a newly trained practitioner with two different training protocols. We evaluated the agreement between them by calculating the weighted kappa of ultrasound severity grades for each quadrant and by the Bland and Altman method to evaluate agreement for GS.

## Results

### - Participants

We included 107 patients with suspected or diagnosed COVID-19 between March 19, 2020 and April 1, 2020 in the EU of the 3 APHP sites and who underwent LU; 107 patients had both LU and chest CT.

### - Descriptive data

Main characteristics of study participants are summarized in Table 2. There were 69 men (64.5%) and the mean age was 61.2±16.6 years (all sites combined). Only one patient was brought in by emergency medical service ambulance. RT-PCR testing for COVID-19 was performed for most (n = 97) patients (10 [9.3%] patients did not have RT-PCR) and 68 (70%) tests were positive. An expert performed LU examinations in 73 (68.2%) patients: 17 (16.7%) were active smokers, 31 (29%) had known hypertension and 17 (15.9%) had type 2 diabetes (Table 2). On admission, oxygen saturation was less than 95% for 50% of patients.

Quantification by CT by an ordinal scale was available for only 90 patients, since the information was missing in the radiologist report..

### - Outcomes data (Table 3, Table 4)

The results obtained in each quadrant are in Table 3.1 and 3.2. For each field (right and left chest walls), the total score ranged from 0 to 12. On the right wall, respectively the left wall, the quadrant most severely affected was the postero-inferior, with 67 patients, respectively 66, showing severity ≥ 2.

For the 107 patients, the mean severity score for all pulmonary quadrants (maximum achievable score: 24) was 9.6± 6.0. Chest CTs were pathological for 101 (94.4%) patients and considered typical of COVID-19 for 86 (85.1%) (Table 4). The severity was minimal (grade 1) for 21 (23.3%) patients and severe (grade 3) for 29 (32.2%).

### - Main results

Considering LU as a 4-category ordinal scale versus CT scale, severity assessment by naive average scores showed only moderate agreement between the 2 scales, as shown by a weighted kappa of 0.52 (95% confidence interval 0.38-0.66) (Figure 1).

**Figure 1.**
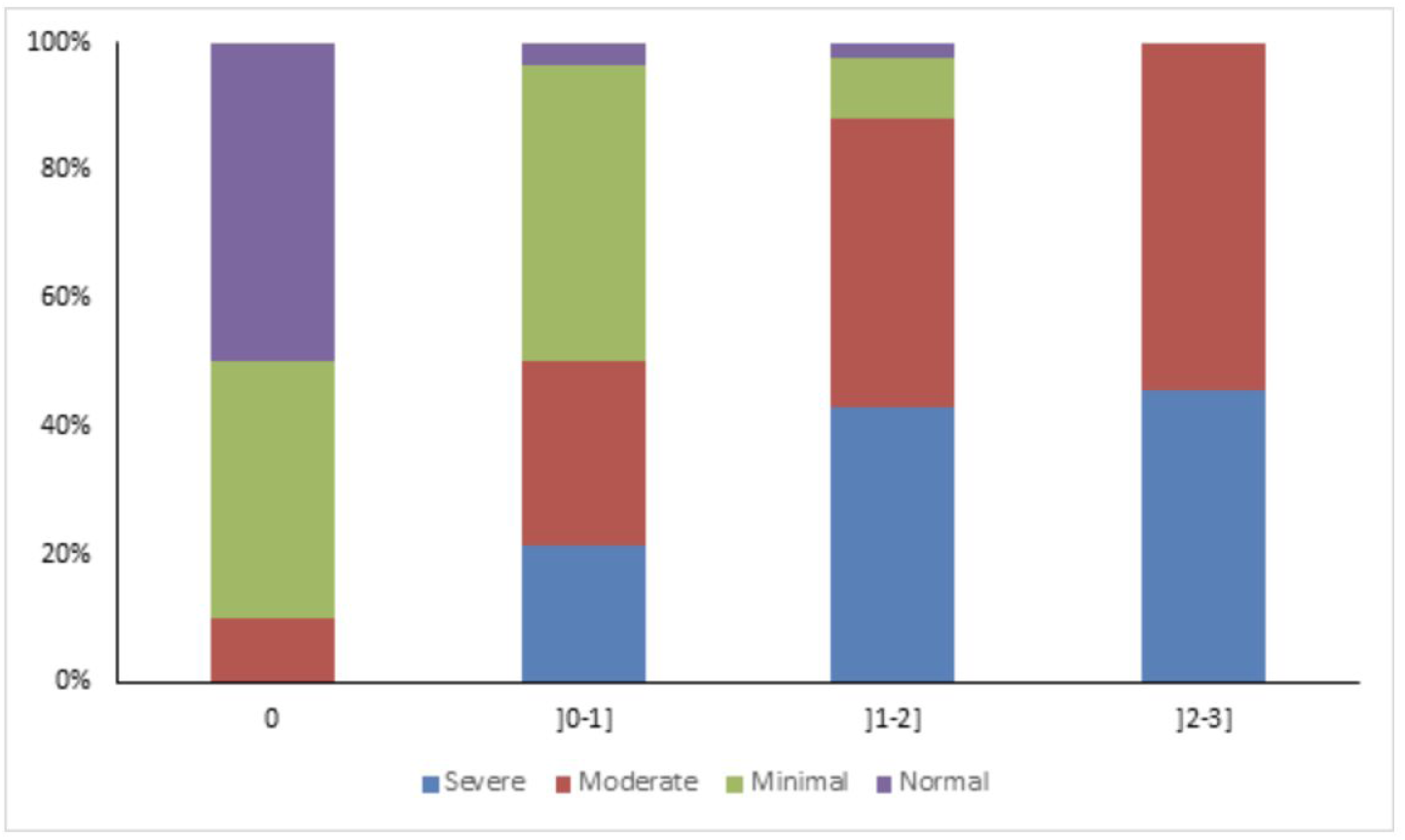
Levels of severity of the disease evaluated by scanner for different level of severity evaluated by echography (Weighted kappa for agreement between level of severity evaluated by echography and scanner χ = 0.54[0.40–0.68]

### LU versus CT score

We found strong relationships between GS and CT evaluation of disease severity (Figure 2). The GS showed good performance to predict evaluation of the disease by CT classified as normal versus pathologic: AUC 0.93 and Brier score 0.04, a value of 1 corresponding to maximal Youden index associated with sensitivity 95% and specificity 83%. This comparison was computed for 90 patients, because 17 had a pathological/normal CT status but missing CT severity score.

**Figure 2.**
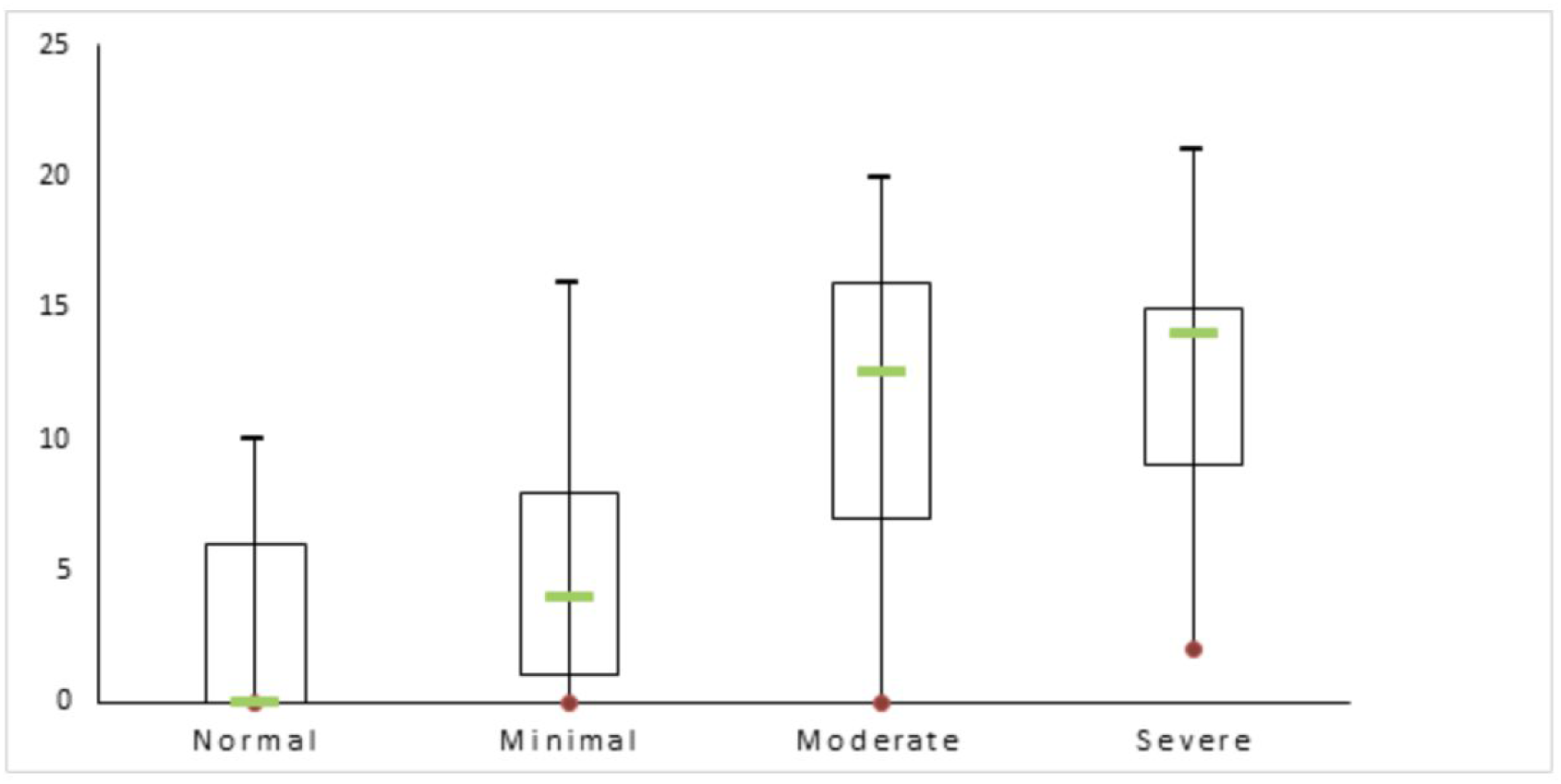
Global echography scores for different severity of disease evaluated by scanner. Box-plot graph shows Median and quartiles (Kruskall-Wall s test, p <0.001)

When a simplified weighted score was built, based on multivariate logistic regression, with SWS equation calculated as 2 x AS-R + 2 x PI-R + 1 x AS-L+3 x PI-L (capital letters correspond to Right, Left, Anterior, Posterior, Superior, Inferior), AUC was slightly higher for prediction (*normal* vs *pathologic*): AUC 0.95 and Brier score 0.0347. A value of 1 was associated with the same sensitivity (95%) and specificity (83%) as the sum of score, but the highest Youden index was obtained for a value of 6 associated with sensitivity 82% and specificity 100%.

In both cases, bootstrap internal validation demonstrated a very small degree of optimism (−0.0025 and −0.0016 for the C-statistic and Brier score for GS, and 0.0018 and −0.0013, respectively, for SWS).

Similar performances were found when CT results were classified as *normal or minimal* versus *moderate or severe:* AUC 0.89 and Brier score 0.12, a value of 7 corresponded to the maximal Youden index associated with sensitivity 86% and specificity 78%. In this case, the SWS equation was: 1 x AS-R + 1x AIL-R + 1 x PS-R + 1 x PI-R + 1 x AS-L+1 x PI-L: AUC 0.90 and Brier score 0.11, a value of 6 corresponding to the maximal Youden index associated with sensitivity 92% and specificity 74%. Here also, a very small degree of optimism was found (−0.024, −0.0153 and 0.0024, −0.0047 for the C-statistic and Brier score for GS and SWS, respectively).

Note also that performance of multivariate logistic regression models and the SWS were very small for both classifications of CT (i.e., differences in AUC were 2.5% and 2%, respectively).

In contrast with previous results, the performance of GS or SWS was lower to predict disease classified as severe by CT (*normal or minimal or moderate* vs severe) (AUC 0.79 and 0.76, respectively).

### Agreement between experts and newly trained physicians (Figure 3, Figure 4)

**Figure 3.**
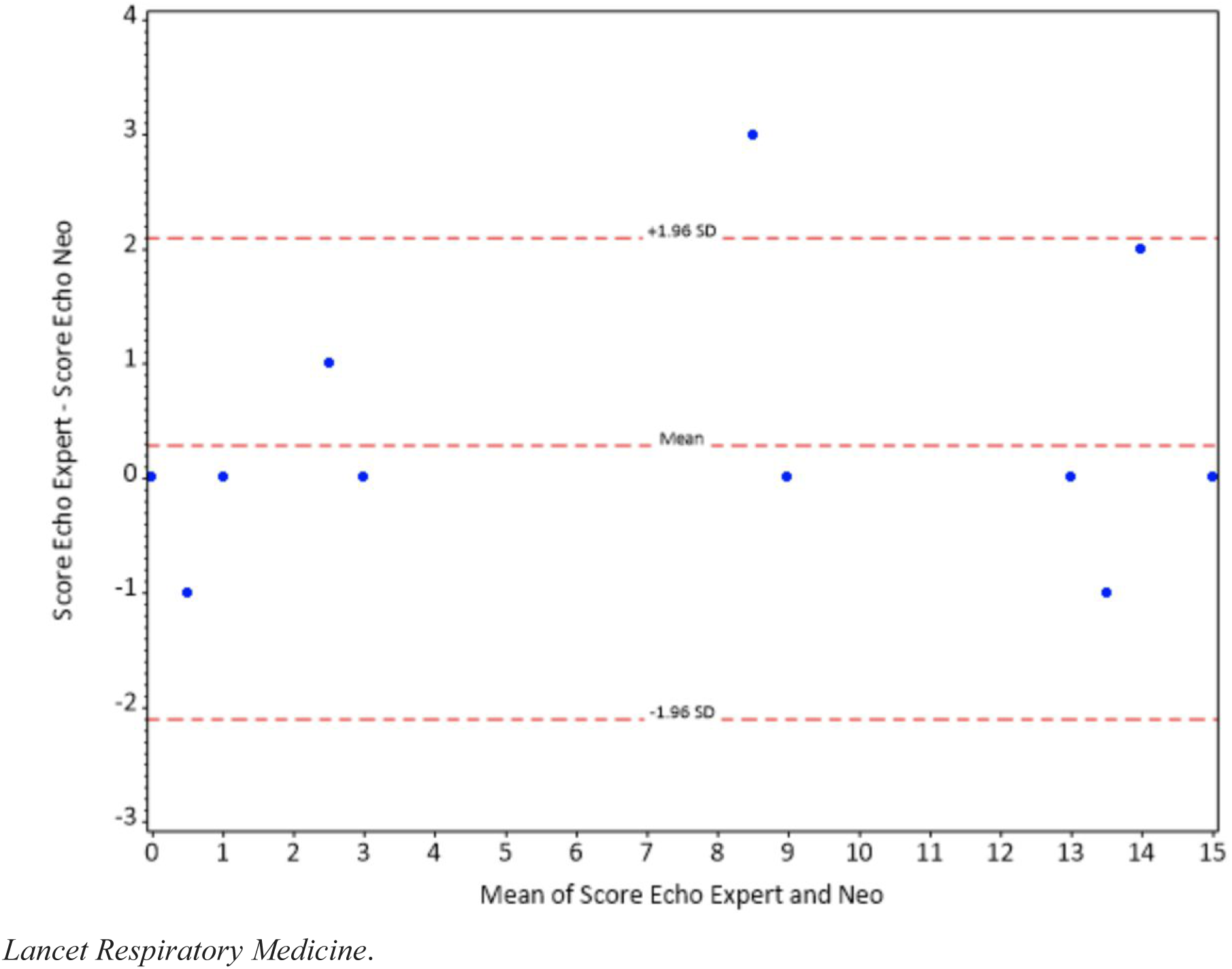
Bland and Altman plot of agreement between global echography scores evaluated by experts and new practitioners (N=14 pairs of raters. some points are confounded for several pairs of raters).

**Figure 4.**
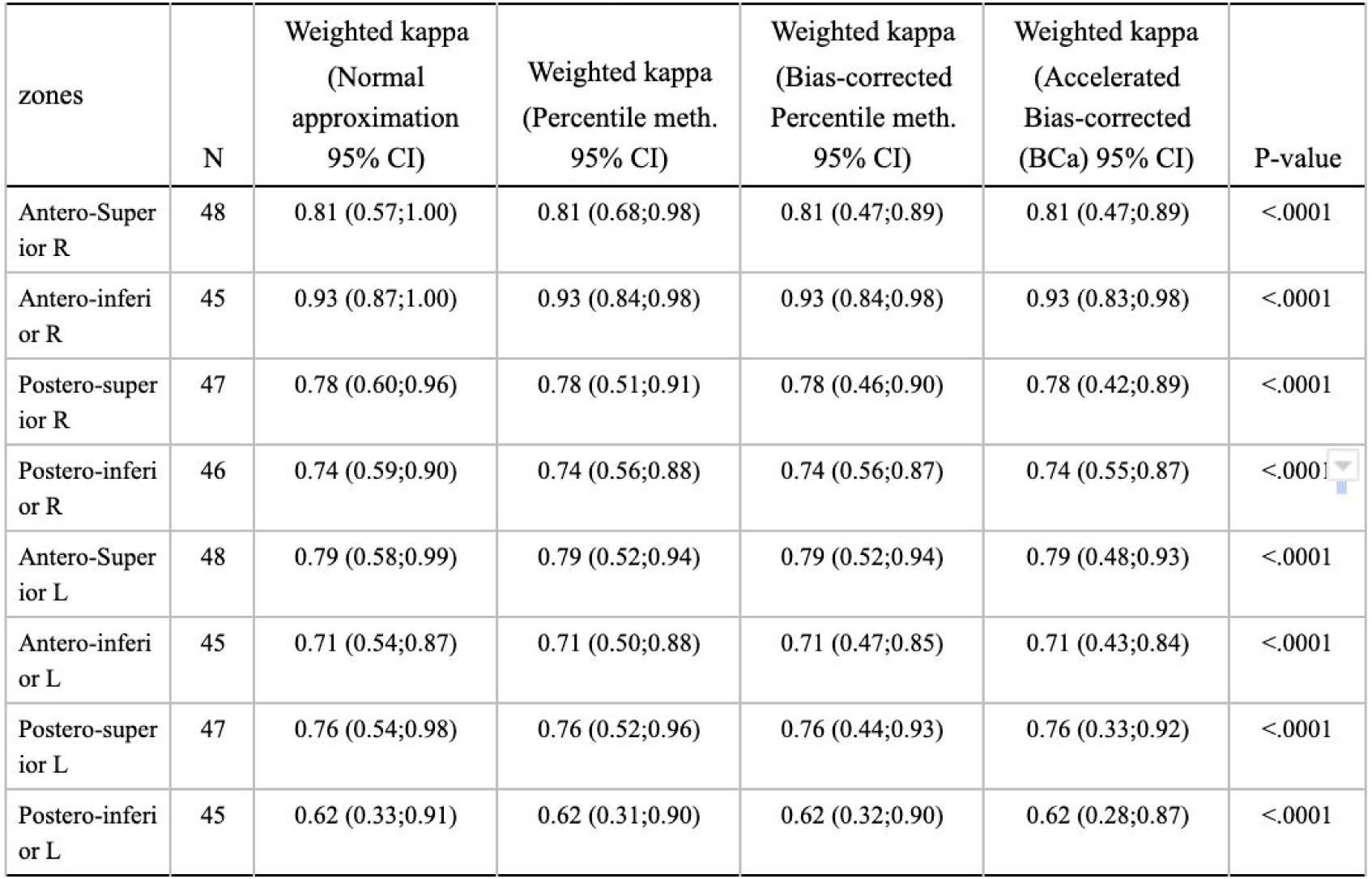
Agreement Expert vs New Trainee

As shown by Bland and Altman plots, we found good agreement between GS evaluated by new trainee with a complete protocole (theory + practice) and expert (n=14 pairs of raters, 1 new trainee). Good agreement was also found when considering each quadrant individually: in all cases, the weighted kappa ranged from 0.85 to 1 (values obtained for PS-L and AS-R, respectively).

When considering all new trainees, we found a moderate agreement considering each quadrant individually (n=48, 4 new trainees), with weighted kappa 0.62-0.81.

## Discussion

In this observational study including 107 patients with suspected or diagnosed COVID-19, LU and chest CT lung damage detection were quite consistent. As for lung damage severity assessment relating GS, consisting of summing the severity of 8 chest points, to CT severity score, defined by extension of lung lesions, we found an AUC of 0.93. Another key, though very preliminary finding, is the concordance in LU scoring between an expert operator and newly trained operator with a complete training protocol (theory + practice), weighted kappa > 0.85. When considering all newly trained physicians, the agreement by chest zones was less satisfactory, weighted kappa 0.62-0.81.

### - Limitations of the study

LU severity score GS may be improved in many ways. First, the score carries information about the severity of lung injuries but does not sufficiently address their extension. For instance, for equal grade 3 scoring, some patients may have only one chest point graded 3 while others may have 3 chest points graded 1. Hence GS does not reflect the disease extension. Scores such as the median, the average or the maximum severity ultrasound grading are not pertinent. They are too naive to include specific or sufficient information on the extension of lung lesions. Besides, let us indicate that we collapsed the CT score “extended”, “severe”, “critical” to “severe” out of clinical simplicity, in order to match the 4 grades ultrasound scoring. In order to improve the ultrasound scoring, a further study is ongoing comparing CT and LU for each chest point, and associated with more refined statistics based on machine learning techniques taking into account linear and non-linearity effects such as the non-linear jump in condition between GS at 0 and 1. Although this approach suggests gains in specificity, a more complex scoring may not be easy to compute in practice.

Only 6 CT were normal. Hence, we lack sufficient data to estimate false positive rate. Also, only 10 ultrasound exams had a 0 grading. Hence, we lack sufficient data to estimate negative predictive value.

Also, we merged results from patients consulting in EUs of Hospital Cochin and Hospital Lariboisière with patients admitted to EU of Hospital Hôtel Dieu converted to a screening unit for APHP medical staff with suspected COVID-19. All included patients were symptomatic but we did not yet investigate the difference in characteristics of patients depending on the centre they were refered to, and patients in the screening unit may have had less severe conditions.

### Interpretation of results

The satisfactory sensitivity of LU examination was not surprising, and authors have already documented this point [11, 16]. One issue is the lack of specificity, so LU findings must be interpreted with caution, especially since the viral syndrome is not specific and should not lead to rule out other causes of dyspnea, especially because patients may have over-added pulmonary embolism [17]. In our study, the clinician in charge of the patient and the LU operator were blinded to each other’s results. Specificity may be improved with interpretation in light of the clinical context. With better specificity, one may hope that LU could help appreciate the more or less typical character of COVID-19 disease and make it not only a screening tool but also an orientation diagnostic tool.

Furthermore, if LU seems to be a reasonable alternative to chest CT in situations of compromised availability, LU must not compete with chest CT, especially when the patient requires closer lung status evaluation. Indeed, patients may not be “ultrasoundable”, (morbid obesity, sub-cutaneous emphysema or any cause that prevents LU interpretation). Also, patients with pre-existing conditions such as emphysema, fibrosis etc. may have an abnormal LU affecting the relevance of the operator’s interpretation. Due to the principle of ultrasound technique lesions without any contact with the pleural envelope will not be detected (air interposition). Although LU may be informative in terms of many conditions such as pneumothorax, pleural effusion, interstitial images or sub-pleural consolidation. CT has remarkable specificity for lung lesions and is the reference imaging for thorough investigations.

There is an abundant literature on ultrasound training protocols. Some evidence suggests that short protocols may be sufficient especially when the training is focused on specific medical issues. [18, 21] This seems in line with our observations. Besides, one might think that completing ultrasound theory training with practice has a positive impact on new trainees performances. However, this observation is strongly limited due to their few number.

### Generalisability of the study results

The results of this study suggest that LU might find a place in management of suspected or diagnosed COVID-19. The point-of-care nature of the exam, the accessibility of the devices (relatively low-cost, handheld), the real-time interpretation and the non-invasive technology may suggest LU as a major screening tool.

At the basic level, with CT availability issues, because of poor health infrastructures or lack of access to this type of resource in the context of significant demands on CT resources, LU may offer the advantage of quick and lightweight screening of patients. A quick skills acquisition curve may help with dissemination of this practice. Moreover, in highly degraded contexts, LU could be a diagnostic tool based on a clinical-radiological correlation reasoning.

We cannot yet give a precise answer for the use of LU in COVID-19 patients follow-up. The authors are currently conducting a multi-centric prospective study, correlating LU results with clinical evolution outcomes measured up to 1 month after the initial LU, which should help provide answers.

One important subject of concern is hygiene and over-risk of operator contamination., The French learned society of radiology emitted a guidelines stating that ultrasound imagery has no proven place in Covid19 patients management [22]. Our approach is to target ultrasound imagery use as a patient’s bedside tool, which has to be carried out by the clinician in charge. One may reasonably expect from the latter the use of a stethoscope, which exposes the operator to the same level risk of contamination. Moreover, we mention this article [23], comparing the safety of ultrasound against stethoscope, and concluding favorably for the ultrasound.

Finally, in our study, although the agreement results between expert and new trainees, are rather satisfactory, training protocols may be improved and tested with a larger pool of newly trained physicians.

## Conclusion

LU allows for assessing the severity of lung injuries with patients suspected or diagnosed COVID-19 and is consistent with chest CT findings. This examination, carried out in a few minutes, at the patient’s bedside, interpretable in real time and by the doctor in charge, with a non-radiation technology, and relatively low cost. represents a timely opportunity to use LU as a triage tool, especially with issues of the availability of CTs because of overwhelming requests, quite common in the COVID-19 pandemic context, or of poor health infrastructure. Moreover, the short learning curve may help spread the practice beyond the ultrasound expert community.

## Data Availability

At this time, the data is still not freely available.

## Authors’ contributions

Dr Mehdi Benchoufi designed and coordinated the study, participated to the statistical analysis plan setting and the construction of the Ultrasound severity assessment score.

Dr Jérôme Bokobza co-designed the study, set the ultrasound examination procedure and built the lung damage ultrasound grading.

Dr Anthony Chauvin set the ultrasound examination procedure, reviewed the study design and the article

Pr Elisabeth Dion set the framework for Chest CT-scan place in the study (relevant informations from radiologist report, lung damage severity score, etc), led the secondary analysis of chest CT-scan for confirmation of radiologists reports and to explore the limits of Ultrasound in terms of specificity.

Dr Marie-Laure Baranne: participated to the design of the study, contributed to statistical analysis plan and reviewed extensively the article.

Dr Fabien Levan participated to the design of the ultrasound examination procedure and the statistical analysis plan setting.

Dr Maxime Gautier, Dr Delphine Cantin, Dr Thomas d’Humières, Dr Cédric Gil-Jardiné, Dr Sylvain Benenati, Dr Mathieu Orbelin, Dr Mikaël Martinez, Dr Nathalie Pierre-Kahn contributed equally. They are all ultrasound experts, with a substantial experience in lung ultrasound imaging. They all brought their expertise to review the study design, to validate inclusion and exclusion criteriae, the lung examination procedure, and the simplified lung severity ultrasound grading. They participated to the discussion section.

Abdourahmane Diallo executed the statistical analysis plan under the supervision of Eric Vicaut.

Pr Eric Vicaut designed the statistical plan, led the analysis, designed the Ultrasound Score by challenging several refined statistical models. He extensively reviewed the study design and brought synthetic insights on opportunities and limits of the ultrasound scoring.

Dr Pierre Bourrier is a radiologist, specialised in Ultrasound imaging. He designed the theoretical framework of the place of quick ultrasound imaging for triage purpose, and in particular for patients suspected of viral lung infections, and helped derived applications in the context of Covid19 pandemics. He co-designed the training protocol. He reviewed the study design.

All the authors finally reviewed the article and approved the article.

## Ethics committee approval

According to french regulatory rules, the study was approved by the ethical committee N°IRB: IORG0009855 (COMITE DE PROTECTION DES PERSONNES DU SUD-OUEST ET OUTRE-MER 4) with N° CPP2020-03-035a / 2020-A00768-31 / 20.03.24.70826

## Aknowledgements

The sponsor of the study is Assistance Publique des Hôpitaux de Paris (APHP). We applied for a grant from the French Ministry of Health. Should our project not be retained, APHP would provide the necessary funds.

